# Awareness and utilization of free maternal healthcare services among women in Mt. Elgon Sub-County, Kenya

**DOI:** 10.1101/2023.01.25.23285004

**Authors:** Oliver Furechi

## Abstract

Maternal mortality remains a matter of public health concern with over 300,000 women dying while giving birth annually. This has driven policy makers to come up with free maternal healthcare services as policy intervention. The Kenyan Government adopted the free maternal healthcare policy in 2013. The policy exempts maternal services from user fees in all public health facilities. This is aimed at promoting skilled delivery to reduce pregnancy-related mortality. In Mt Elgon sub-county, only 45 percent of women deliver in a health facility, this is below the national average of 61.5 percent.

This paper presents results from a cross-sectional study conducted in January 2021 in Mt Elgon Sub-County among 377 randomly selected women who delivered in the preceding 12 months and 4 FGDs with women of reproductive age. Descriptive statistics were undertaken while Chi-Square test was used for inferential statistics. Andersen behavioral model of health service use was used to examine how womens demographic characteristics and awareness regarding the free maternal health policy has affected utilization of maternal healthcare services. The results indicate that respondents were generally aware that there is a government policy on free maternal health policy. However, gaps still exist in terms of pushing awareness towards universal reach, infrastructure development, health worker patient ratio, timely provision of essential supplies and long distance to health facilities This calls for increased sensitization utilizing community health volunteers and allocation of financial and human resources allocation to the department of health within County Governments.

## INTRODUCTION

Maternal mortality remains a matter of public health concern. The world health organization (WHO) estimates that globally 300,000 women die each year with over 90 percent living in low-middle income countries[1]. These preventable mortalities are occasioned by several factors hindering women from accessing and utilizing maternal healthcare services[2] moreover, beyond mortalities, many more women suffer from short and long term disabilities and illness associated with pregnancy such as depression, infertility and fistula[3] According to WHO, the major causes of maternal deaths comprise; hemorrhage (excessive loss of blood), hypertensive disorders and sepsis/infection which account for more than half of maternal deaths[4]. Most of these deaths could be averted through access to care during pregnancy and delivery period[5]. Access and utilization of antenatal care, skilled birth attendance and postnatal care are therefore critical in reducing maternal mortalities [6,7].

In Kenya, the period 2000 - 2015 saw little or no progress towards achievement of MDG 5 on reducing maternal mortalities. Maternal mortalities remained high at 362 deaths per 100000 live births, this fell short of the 147/100,000 target [8]. It is against this backdrop that in 2013, the Kenyan Government instituted free maternal health care service to encourage women to take up antenatal, intrapartum, and postnatal healthcare services. The free maternal healthcare service policy is commonly referred to as *Linda mama program*, which means protect women. The Linda mama program reimburses health facilities based on their capacity to manage pregnancy and delivery complications. As such, 2500 ($25) Kenya shillings are reimbursed for every delivery conducted in level 2 facilities (health centers) and level 3 health facilities (sub district hospitals); 5000 ($50) Kenya shillings are reimbursed for every delivery carried out in level 4 health facilities (district hospitals) and level 5 health facilities (provincial hospitals); and Ksh 17,500 ($175)is reimbursed for every delivery performed in national referral health facilities[9]. The 2014 Kenya demographic health survey[10] indicates that over 90 percent of women attended at least one antenatal care clinic compared to only 58% that completed the recommended four visits. Moreover, only 62 percent of women deliver with a skilled healthcare provider. Studies [9–12] have identified demographic factors that determine utilization of healthcare service with age, education, socio-economic status, marital status and place of residence being significant factors. Additionally, the level of women’s awareness of available health services and associated costs has an important bearing in determining utilization of maternal healthcare services [15]. A study in India [16] found that 98% of respondents were aware of different types of family planning methods but only 68% used some family planning method. Furthermore, in Ghana mothers with low knowledge levels on danger signs in pregnancy were less likely to use postnatal care services [17]. While in Uganda utilization of maternal health services varied greatly based on demographic and economic factors of women with those living far away from towns and with low levels of awareness having low levels of utilization[18]. The studies conducted in Kenya after initiation of free maternal healthcare services focused on implementation of the policy and utilization of the services without considering awareness as a factor that influences utilization[10], [20–22], moreover, within the country disparities in utilization exists[23,24] thus there is need for more nuanced understanding at the local level to inform decisions the newly created County Governments as per the 2010 Constitution. An understanding of such determinants is critical in aiding planning and delivery of maternal healthcare services that is accessible to local communities to improve health outcomes.

## METHODS

We conducted a mixed method cross-sectional study that examined how awareness influences utilization of free maternal healthcare services among women. The study was carried out between January and March 2021 in Mt. Elgon Sub County within the larger Bungoma County. The area is part of western Kenya bordering Uganda to the west. It is mountainous and largely inhabited by the Sabaoti ethnic group. The road network is poor with access to some areas using a motorised vehicle virtually impossible. *Boda-boda*^1^ and donkey carts are the common means of transport. The Sub County has 12 health facilities: 10 dispensaries, one sub county hospital and one faith based health facility. These facilities serve a total population of approximately 65,402. An average of 6200 women deliver in any given year, and 45% of these women deliver within health facilities[10].

The primary study population was 6,327 women who had delivered in the preceding 12 months. Using Yamane (1967) formula a sample of 377 was included in the study. Simple random sampling technique used to identify respondents from a list of 6,327 extracted from community-based health information system (CBHIS). Community health Volunteers (CHVs) guided research assistants to households where selected women stay and did the initial introduction and allowed the research assistants to proceed to administer the semi-structured questionnaire in a private place. Through purposive sampling 5 health managers and 8 CHVs were identified and interviewed as key informants. Additionally, we conducted two FGDs with women who were pregnant or lactating. The semi-structured questionnaire and FGD tools were pretested in a nearby subcounty with similar characteristics to the study area, the tools were revised based on the feedback received.

The independent variable was awareness of free maternal healthcare services while utilization of antenatal service and delivery with skilled birth attendant were the dependent variables. The study was anchored on behavioral model of health use developed by Andersen[24]. In this framework, individuals’ level of awareness is considered as a predisposing factor.

Using Social Package for Social Sciences version 18 quantitative data was analysed to generate descriptive statistics; frequencies, means, percentages while Chi square was used to determine the relationship between independent and dependent variables. P<0.05 within 95 percent confidence interval was considered significant. Qualitative data was thematically analyzed and reported as verbatim quotes. The study was approved by Maseno University Ethics and Research Committee Approval No. MUERC/00880/20. The Mt. Elgon Sub County health management team allowed the study to be undertaken within their jurisdiction. Study respondents were informed of the objectives of the study and their right to participate voluntarily prior to providing written consent.

## RESULTS

### Socio-demographic and economic characteristics of the respondents

A total of 377 respondents were interviewed. Most of the respondents 208(55.2%) were within the age bracket 26-34 years, while those within the age group 18-25 were the least represented at 26 (6.9%). In terms of level of education, 251(66.6%) had achieved primary level education with a quarter (24.9%) having attained secondary education, a paltry 4(1.1%) had completed degree level education. Analysis of socio-economic status revealed that more than two-thirds 256(68%) of the respondents belonged to the first and second quintile. A paltry 4.8% (18) were in the fifth quintile see Table 1 for more details.

**Table 1:**
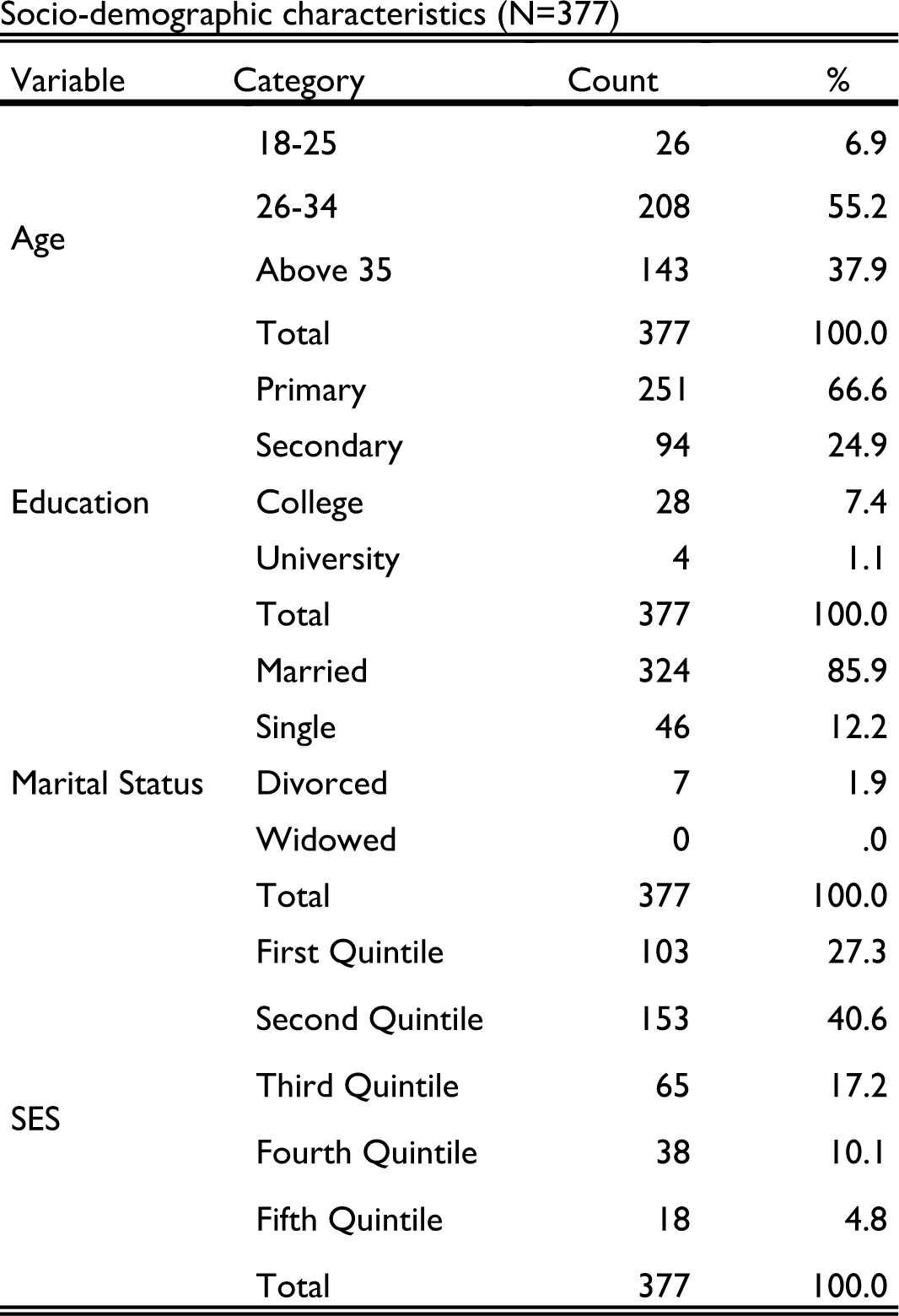
Socio-Demographic and Economic Characteristics of Respondents

| Variable | Category | Count | % |
| --- | --- | --- | --- |
| Age | 18-25 | 26 | 6.9 |
|  | 26-34 | 208 | 55.2 |
|  | Above 35 | 143 | 37.9 |
|  | Total | 377 | 100.0 |
| Education | Primary | 251 | 66.6 |
|  | Secondary | 94 | 24.9 |
|  | College | 28 | 7.4 |
|  | University | 4 | 1.1 |
|  | Total | 377 | 100.0 |
| Marital Status | Married | 324 | 85.9 |
|  | Single | 46 | 12.2 |
|  | Divorced | 7 | 1.9 |
|  | Widowed | 0 | .0 |
|  | Total | 377 | 100.0 |
| SES | First Quintile | 103 | 27.3 |
|  | Second Quintile | 153 | 40.6 |
|  | Third Quintile | 65 | 17.2 |
|  | Fourth Quintile | 38 | 10.1 |
|  | Fifth Quintile | 18 | 4.8 |
|  | Total | 377 | 100.0 |

### Awareness of free maternal healthcare services

A total of 248 (65.8%) were aware of the free maternal health care services. Further analysis revealed that out of the 129 (34.2.%) who were not aware, two thirds (68.2%) of them had primary level education, were relatively younger (26-34) and unemployed. A community health volunteer averred. *“most of these young girls who get pregnant for the first time are the ones whom we now focus on more to help them understand motherhood and enable them access critical information they need to know”*.

Furthermore, an analysis based on respondent’s social economic status(SES) indicates that all the 18 respondents in the fifth quintile were aware of the existence of free maternal health care services compared to 95 (92.2%) of the respondents within the first quintile. Although the association between SES and awareness was not significant X^2^ (4, *N*=377) = 7.732 p>.05. To support the high levels of awareness, A woman in the FGD said,

*“we just hear of Linda mama (the name for free maternal health services) from community health volunteers who encourage us to come to health facilities as it is free…we are told it is free”*

Age of the respondent was not a significant factor determining level of awareness of free maternal health care services X^2^ (2, *N*=377) = 3.647, P>.05. However, education was significantly associated with awareness X^2^ (3, *N*=377) =8.023, P<.05. with all respondents with College or University education being aware of free maternal healthcare services. Moreover, finding indicate that there is no significant association between socioeconomic status with awareness X^2^ (4, *N*=377) =7.732, P>.05).

Awareness of specific entitlements provided within the free maternal health care service policy was low. Only half 132 (53.2%) of those who were aware of the free maternal health care services mentioned all the four services, which includes: Antenatal profiling, four antenatal care visit, skilled delivery, and postnatal care. ANC profiling was the least mentioned service entitlement. Even community health volunteers seem not to understand the free maternal healthcare services entitlements very well.

*“You know sometimes they* (healthcare workers) *ask women to pay for the lab…in other cases they don’t…but we usually tell women as part of their preparation to also set aside some money for emergency”*. A CHV participant in KII.

Moreover, service providers also say the policy is not clear as it does not itemize what it covers, this leads to confusion. One health manager who participated in KII said

*“A woman can come to the hospital, she is pregnant but on diagnosis she is suffering from non-pregnancy related disease to her she knows she shouldn’t pay because it is free……we need a clear guidance and sensitization of community members on what is covered and what is not covered…*..*some health facilities charge women for ANC profiling* (Lab services) *while others don’t…*.*it is not clear”*.

### Awareness and uptake of antenatal and skilled birth attendance

Studies [27]–[29] have indicated that there is an association between awareness of healthcare services and utilization. In this study, a cross tabulation of awareness and utilization of four antenatal care services ((ANC4) indicate that those who were aware of free maternal health services were more (65.8%) likely to complete ANC 4 as compared to those who were not aware(34.2%) see table 2 for details. Results of Chi square test of independence indicate that awareness was significantly associated with utilization of four ANC clinics X^2^ (1, N=377) =4.85 p<.05.

**Table 2:**
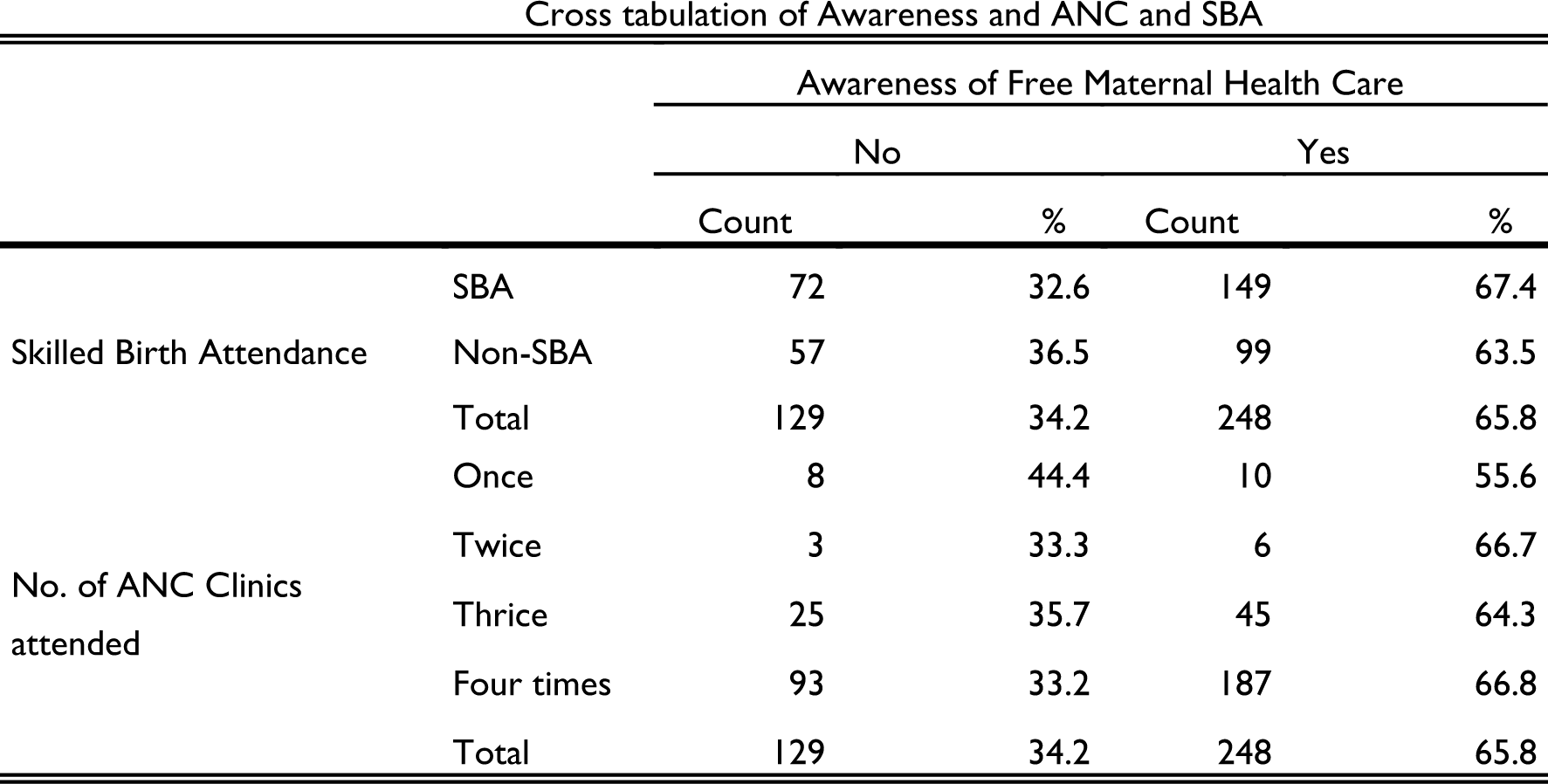
Awareness and utilization of Antenatal Care and Skilled Birth Attendance services.

Additionally, attendance of four antenatal care services increases the possibility of delivering in a health facility with a skilled birth attendant. Majority of the respondents 280(74.3%) reported four antenatal care visits while only 27(7.2%) managed to attend one or two clinics during their last pregnancy. A woman who participated in FGD said. *“It is important to go even for once so that you can be issued with the booklet* {mother baby booklet} *so that you are safe and able to use it during delivery and immunization of the baby not to forget the bed net”*.

## DISCUSSION

More than 45% of the respondents were not aware of the free maternal healthcare services. This implies that women in this setting are likely to have missed opportunities for critical live saving maternal healthcare services [33]. Unlike the findings Yaya et al,. (2017) in their study in Bangladesh, the level of education, socio-economic status was not significantly associated with awareness. Similar findings by are highlighted by Chen et al [30] who concludes that awareness is an important factor that determine demand for preventive medical care among the elderly. Furthermore, Rutaremwa et al., (2015) had a similar finding on education this study differs on their finding on association socioeconomic status with awareness which was not significant Additionally, Utilization of healthcare services is influenced by the ability of households to cater for the associated costs[34]. In this study, an understanding of household social economic status (SES) unraveled that those in higher socio-economic status have a higher rate of utilization of antenatal care and skilled delivery as compared to the poor. This results are similar to other studies [19–22] which concluded that despite removal of user fees access to healthcare is highly dependent on individual’s socio-economic status.

The younger cohort in this study (18-25) were fewer among those who completed the recommended four antenatal care visit and delivered with a skilled health provider, these findings reflect the general understanding of young people’s uptake of reproductive health services which is influenced by cognitive and psychosocial barriers. The cognitive barriers are related to awareness of maternal knowledge and available services [36].

Likewise those above 35 years also had depressed uptake of maternal services, this can be interpreted to imply that women with more than one birth are less likely to use maternal health, this assertion is supported by a reanalysis of demographic health survey data by Mbugua and MacQuarrie 2018 ([37] who found that women above forty years have less odds ratio as compared to those within 20-29 years bracket. This demonstrates that it is important to focus awareness creation among women who are pregnant for the first time.

Unlike the findings of Gitimu et al.,2015[38] who concluded that uptake of antenatal care and skilled birth attendance increases as the level of a woman’s level of education increases, we found that this situation only applies to those with primary (57.4%) and secondary level (63.8%) education and dropped among those with college/university education (51.8%) in this study.

The implication of low uptake is that women are missing the critical services provided during the four antenatal care visits even health facilities access funds for services not rendered to mothers as the payment is lumpsum and pegged on deliveries reported.

Similar to Ghana’s free maternal health policy which was also introduced as a result of poor maternal and neonatal health outcomes and through a political directive that was later operationalized[32], the level of uptake of antenatal care and skilled birth attendance is on the rise[19]. A sustained upward trajectory in uptake of this critical maternal health services will go a long way in accelerating traction towards achievement of SDG 3.

## CONCLUSION

There was moderate level of women’s awareness of free maternal healthcare services that resulted in depressed utilization. Based on these findings, there is need to improve on awareness creation interventions targeting the younger cohorts within the reproductive age. The existing community health volunteers’ structures and strengthened facility-based health talks will make a huge difference. Beyond the health system, improving the socio-economic status of the people through programs that target poverty reduction will also have a direct effect on maternal mortalities.

## Data Availability

Anonymized data is available from the corresponding author on reasonable request

## Acknowledgements

I acknowledge all the respondents who provided valuable insights including health facility managers and community health volunteers. I appreciate Drs. Fred Ikanda and Mary Ochieng’ who provided guidance for my Master of Arts in Social Policy at Maseno University.

## Footnotes

1 A motorcycle which is hired for transportation.

## Notes

### Competing Interest Statement

The authors have declared no competing interest.

### Funding Statement

This study did not receive any funding

### Author Declarations

An approval was provided by Maseno University Research and Ethic Committee

